# Quantitative SARS-CoV-2 anti-spike responses to Pfizer-BioNTech and Oxford-AstraZeneca vaccines by previous infection status

**DOI:** 10.1101/2021.03.21.21254061

**Authors:** David W Eyre, Sheila F Lumley, Jia Wei, Stuart Cox, Tim James, Anita Justice, Gerald Jesuthasan, Denise O’Donnell, Alison Howarth, Stephanie B Hatch, Brian D Marsden, E Yvonne Jones, David I Stuart, Daniel Ebner, Sarah Hoosdally, Derrick W Crook, Tim EA Peto, Timothy M Walker, Nicole E Stoesser, Philippa C Matthews, Koen B Pouwels, A Sarah Walker, Katie Jeffery

## Abstract

**Objectives:** We investigate determinants of SARS-CoV-2 anti-spike IgG responses in healthcare workers (HCWs) following one or two doses of Pfizer-BioNTech or Oxford-AstraZeneca vaccines.

**Methods:** HCWs participating in regular SARS-CoV-2 PCR and antibody testing were invited for serological testing prior to first and second vaccination, and 4 weeks post-vaccination if receiving a 12-week dosing interval. Quantitative post-vaccination anti-spike antibody responses were measured using the Abbott SARS-CoV-2 IgG II Quant assay (detection threshold: ≥50 AU/ml). We used multivariable logistic regression to identify predictors of seropositivity and generalised additive models to track antibody responses over time.

**Results:** Vaccine uptake was 80%, but less in lower-paid roles and Black, south Asian and minority ethnic groups. 3570/3610(98.9%) HCWs were seropositive >14 days post-first vaccination and prior to second vaccination, 2706/2720(99.5%) after Pfizer-BioNTech and 864/890(97.1%) following Oxford-AstraZeneca vaccines. Previously infected and younger HCWs were more likely to test seropositive post-first vaccination, with no evidence of differences by sex or ethnicity. All 470 HCWs tested >14 days after second vaccine were seropositive. Quantitative antibody responses were higher after previous infection: median(IQR) >21 days post-first Pfizer-BioNTech 14,604(7644-22,291) AU/ml vs. 1028(564-1985) AU/ml without prior infection (p<0.001). Oxford-AstraZeneca vaccine recipients had lower readings post-first dose compared to Pfizer-BioNTech, with and without previous infection, 10,095(5354-17,096) and 435(203-962) AU/ml respectively (both p<0.001 vs. Pfizer-BioNTech). Antibody responses post-second vaccination were similar to those after prior infection and one vaccine dose.

**Conclusions:** Vaccination leads to detectable anti-spike antibodies in nearly all adult HCWs. Whether differences in response impact vaccine efficacy needs further study.

## Introduction

As vaccines against SARS-CoV-2 are rolled out globally, individuals and their clinicians may wish to seek reassurance that vaccination has been “effective” and to understand how long protection is likely to last. Given current vaccines generate an immune response to viral spike antigens, anti-spike antibody titres, associated with neutralising activity,^1–3^ provide a potential surrogate marker of protection. Therefore, understanding the assay- and time-dependent dynamics of post-vaccine anti-spike antibody measurements, how they differ between individuals e.g., by age, gender, ethnicity, and with comorbidities, and how these findings relate to protection, is increasingly important.

Multiple vaccines have been developed globally. In the UK, three vaccines have been approved for use,^4^ with Pfizer-BioNTech BNT162b2 and Oxford-AstraZeneca ChAdOx1 nCoV-19 (AZD1222) the most widely used, with many individuals to date receiving only one dose following an extension of the dosing interval to 12 weeks to maximise initial population coverage. The dynamics and magnitude of the immune response seen in vaccine immunogenicity trials are assay-dependent, predominantly focus on individuals without previous SARS-CoV-2 infection, and use in-house assays developed early in the pandemic, rather than the commercially-available, validated assays now accessible to diagnostic laboratories.^1,5–8^ Emerging real-world data show that nearly all individuals vaccinated with the mRNA vaccines Pfizer-BioNTech BNT162b2 and Moderna mRNA-1273 seroconvert by 21 days post-first vaccine dose, with more rapid seroconversion and higher antibody titres seen in individuals previously infected with SARS-CoV-2 (using in-house ELISAs, and 2 commercial platforms).^9–12^ Fewer immunogenicity data or comparative data for the Oxford-AstraZeneca vaccine are available outside of clinical trials. No trials have published data about whether measured immune markers correspond to observed vaccine efficacy (i.e. protection from infection, hospitalisation or death).

Here we compare anti-spike IgG responses, using a widely-available commercial assay, in healthcare workers (HCWs) following one or two vaccine doses and with the Pfizer-BioNTech or Oxford-AstraZeneca vaccines. We also assess how responses vary between those with and without previous evidence of infection.

## Methods

### Setting

Post-vaccine antibody responses were studied in HCWs from Oxford University Hospitals (OUH), four teaching hospitals in Oxfordshire, UK. Data on previous infections were available from symptomatic testing offered to HCWs with new persistent cough, fever ≥37.8°C or anosmia/ageusia from 27-March-2020 by OUH and from community-based PCR-positive test results shared with the hospital by public health agencies and HCWs. In addition, asymptomatic HCWs were offered voluntary nasal and oropharyngeal swab PCR testing every two weeks and serological testing every two months from 23-April-2020.^13–15^ Staff were encouraged to attend for serological testing prior to first and second vaccination, and additionally around 4 weeks post-first vaccination where the second vaccine dose was due to be given after 12 weeks.

We assessed vaccine uptake rates in HCWs registered for symptomatic/asymptomatic staff testing, which represented the majority of the ∼13,500 staff working for OUH. To avoid including HCWs who had left OUH’s employment before vaccine deployment, we only considered staff who participated in asymptomatic screening, symptomatic testing or vaccination from 01-September-2020 onwards.

The staff vaccination programme began on 8-December-2020, starting with the Pfizer-BioNTech BNT162b2 vaccine, with Oxford-AstraZeneca ChAdOx1 nCoV-19 added from 4-January-2021 and predominately provided to all staff at one acute hospital. Some HCWs received the Oxford-AstraZeneca vaccine in clinical trials beginning 23-April-2020 and were included following unblinding if receiving active vaccine.

### Laboratory assays

PCR tests were performed by OUH and community test centres using a range of assays (see Supplement). Post vaccination anti-spike IgG responses were assessed using the Abbott SARS-CoV-2 IgG II Quant antibody test targeting the spike receptor binding domain (RBD), with results available up to 11-March-2021. The assay cut-off is ≥50 AU/ml, with linear quantification of detected results from 50 to 40,000 AU/ml reported by the manufacturer (and confirmed by serial dilution of monoclonal antibodies, Figure S1).

Pre-vaccination antibody status was assessed using the Abbott anti-nucleocapsid IgG assay (defining readings of ≥1.4 as detected), the Abbott anti-spike IgG, and an anti-trimeric spike IgG ELISA^16^ (detected: ≥8 million units).

### Statistical analysis

Staff were grouped into those with evidence of prior infection, i.e. any positive anti-spike or anti-nucleocapsid antibody test or positive PCR prior to first vaccination, and those without (including staff with no previous serology or PCR testing). Proportions anti-spike positive were estimated by week post-first vaccination, censoring follow-up at the second vaccination, and by week post-second vaccination.

We used multivariable logistic regression to identify predictors of any positive anti-spike antibody result ≥15 days post-first vaccination (but before a second vaccination), considering the vaccine given, previous infection status, age, sex, and ethnicity. We modelled quantitative antibody titres by day since first and second vaccination using generalised additive models, adjusting for age and fitting separate models by vaccine and prior infection status (details in the Supplement).

### Ethics statement

Deidentified data were obtained from the Infections in Oxfordshire Research Database which has generic Research Ethics Committee, Health Research Authority and Confidentiality Advisory Group approvals (19/SC/0403, 19/CAG/0144).

## Results

Overall, 8866/11016 (80%) eligible HCWs were vaccinated. Demographic and occupational details of those vaccinated, and the availability of subsequent serological data are given in Table 1.

**Table 1.**
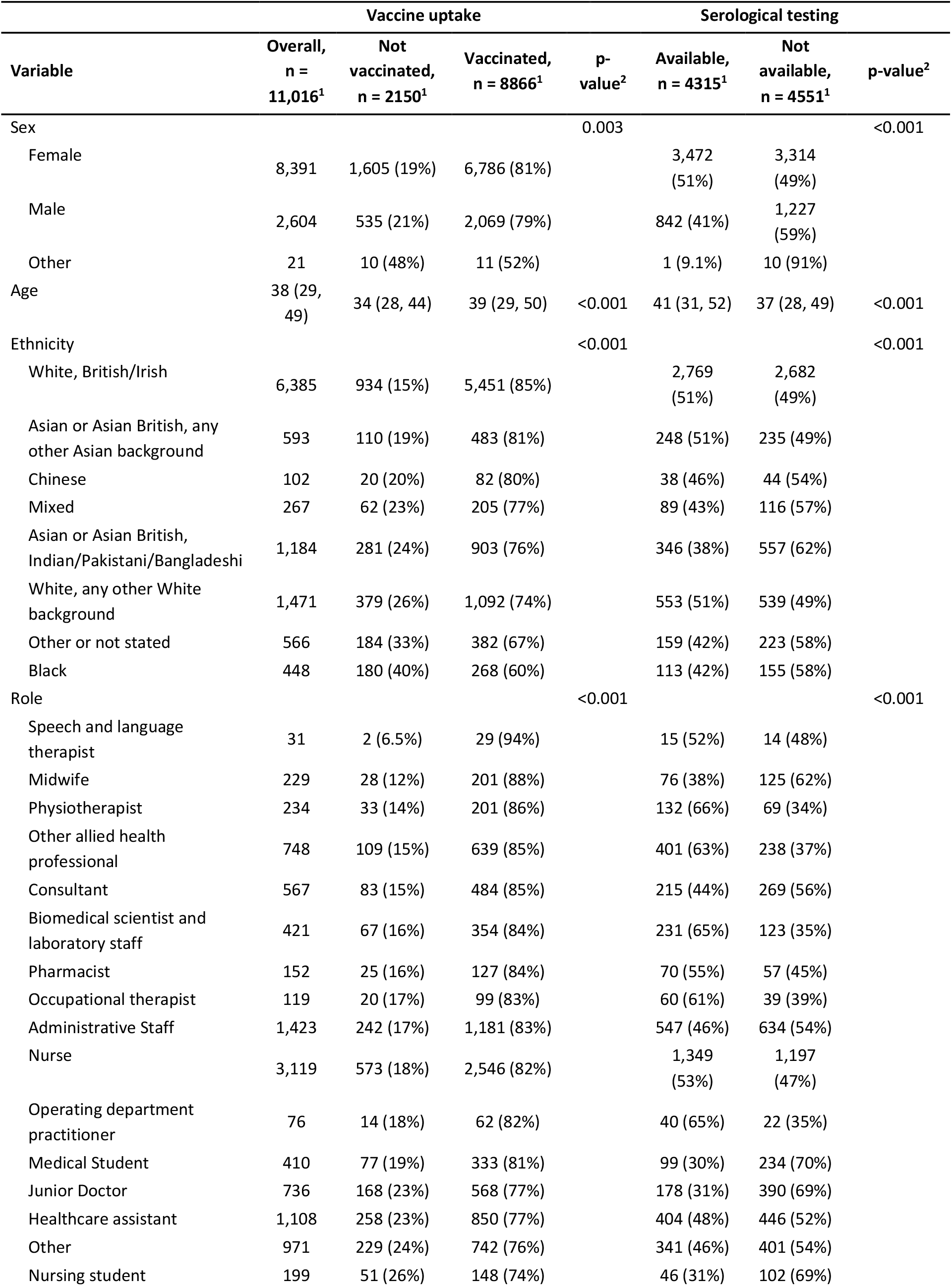
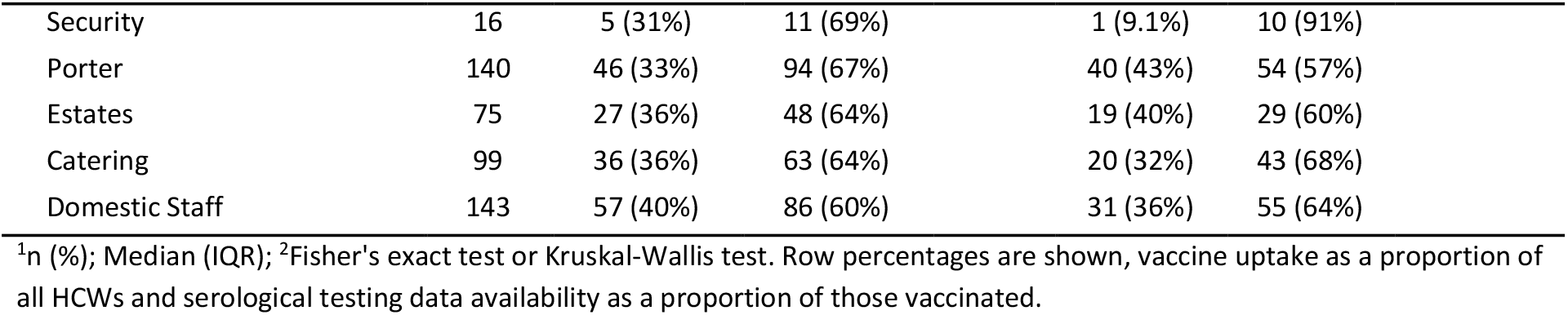
Vaccine uptake rates in healthcare workers by sex, age, ethnicity, and role.

**Table 2.** Factors associated with seropositivity ≥15 days post-first vaccination: univariable and multivariable logistic regression. Test results obtained after a second vaccination are not included. Those of “Other” ethnicity or non-disclosed sex are omitted from the regression models as all tested seropositive.

| Variable | Summary |  | Univariable |  |  | Multivariable |  |  |
| --- | --- | --- | --- | --- | --- | --- | --- | --- |
|  | Anti-spike IgG not detected, n = 40<sup>1</sup> | Anti-spike IgG detected, n = 3570<sup>1</sup> | Unadjusted odds ratio | 95% CI<sup>2</sup> | p-value | Adjusted odds ratio | 95% CI<sup>2</sup> | p-value |
| Previous infection |  |  |  |  |  |  |  |  |
| No prior evidence of infection | 39 (1.3%) | 3,070 (99%) | — | — |  | — | — |  |
| Prior PCR or antibody positive | 1 (0.2%) | 500 (100%) | 6.22 | 0.85, 45.4 | 0.071 | 6.99 | 0.95, 51.3 | 0.056 |
| Vaccine |  |  |  |  |  |  |  |  |
| Pfizer-BioNTech | 14 (0.5%) | 2,706 (99%) | — | — |  | — | — |  |
| Oxford-AstraZeneca | 26 (2.9%) | 864 (97%) | 0.17 | 0.09, 0.33 | <0.001 | 0.17 | 0.09, 0.33 | <0.001 |
| Sex |  |  |  |  |  |  |  |  |
| Female | 33 (1.1%) | 2,921 (99%) | — | — |  | — | — |  |
| Male | 7 (1.1%) | 648 (99%) | 1.01 | 0.44, 2.28 | 0.99 | 0.77 | 0.33, 1.79 | 0.55 |
| Prefer not to say | 0 (0%) | 1 (100%) |  |  |  |  |  |  |
| Ethnic group |  |  |  |  |  |  |  |  |
| White | 37 (1.3%) | 2,774 (99%) | — | — |  | — | — |  |
| Asian | 2 (0.4%) | 480 (100%) | 3.20 | 0.77, 13.3 | 0.11 | 2.50 | 0.59, 10.6 | 0.21 |
| Black | 1 (1.1%) | 91 (99%) | 1.21 | 0.16, 8.95 | 0.85 | 1.34 | 0.18, 10.0 | 0.78 |
| Other | 0 (0%) | 225 (100%) |  |  |  |  |  |  |
| Age | 49 (39, 58) | 41 (30, 51) |  |  |  |  |  |  |
| Age, per 10 year older |  |  | 0.64 | 0.49, 0.83 | <0.001 | 0.66 | 0.51, 0.86 | 0.002 |

<sup>1</sup>n (%); Median (IQR); <sup>2</sup>CI = Confidence Interval

Vaccination rates were slightly higher in women (6786/8391, 81%) than men (2069/2604, 79%; p=0.003). The median (IQR) age was higher in those vaccinated than not (39 (29-50) vs. 34 (28-44) years, p<0.001). Vaccination rates also varied by ethnicity with the highest rates in those identifying as white British/Irish (85%) and as “other Asian background” (mostly Pilipino in our setting, 81%) with the lowest rates in those of Black (60%) or “Other” (67%) ethnicity. Senior doctors (85%), therapists (83-94%) and pharmacists (84%) had some of the highest rates of vaccination, with lower rates in support staff including security, porters, estates, catering and domestic staff (60-69%).

Post vaccine quantitative anti-spike results were available following either a first or second dose or both for 4315 HCWs. 3377 antibody measurements were available (in 2863 HCWs) following a first dose of Pfizer-BioNTech and 1108 (992 HCWs) following a first dose of Oxford-AstraZeneca vaccine. 560 (483 HCWs) and 25 (21 HCWs) anti-spike results were available following a second dose of each vaccine respectively (median (IQR) dosing interval 24 (21-28) days).

### Antibody positivity post-first and second vaccination

Anti-spike antibody responses rose in the 14 days post-first vaccination, such that from day 15 onward near 100% seroconversion was seen regardless of vaccine received or previous infection status (Figure 1). Overall, 3570/3610 (98.9%) HCWs were seropositive when tested >14 days post-first vaccination and prior to second vaccination, 2706/2720 (99.5%) of those receiving the Pfizer-BioNTech vaccine vs. 864/890 (97.1%) receiving the Oxford-AstraZeneca vaccine.

**Figure 1.**
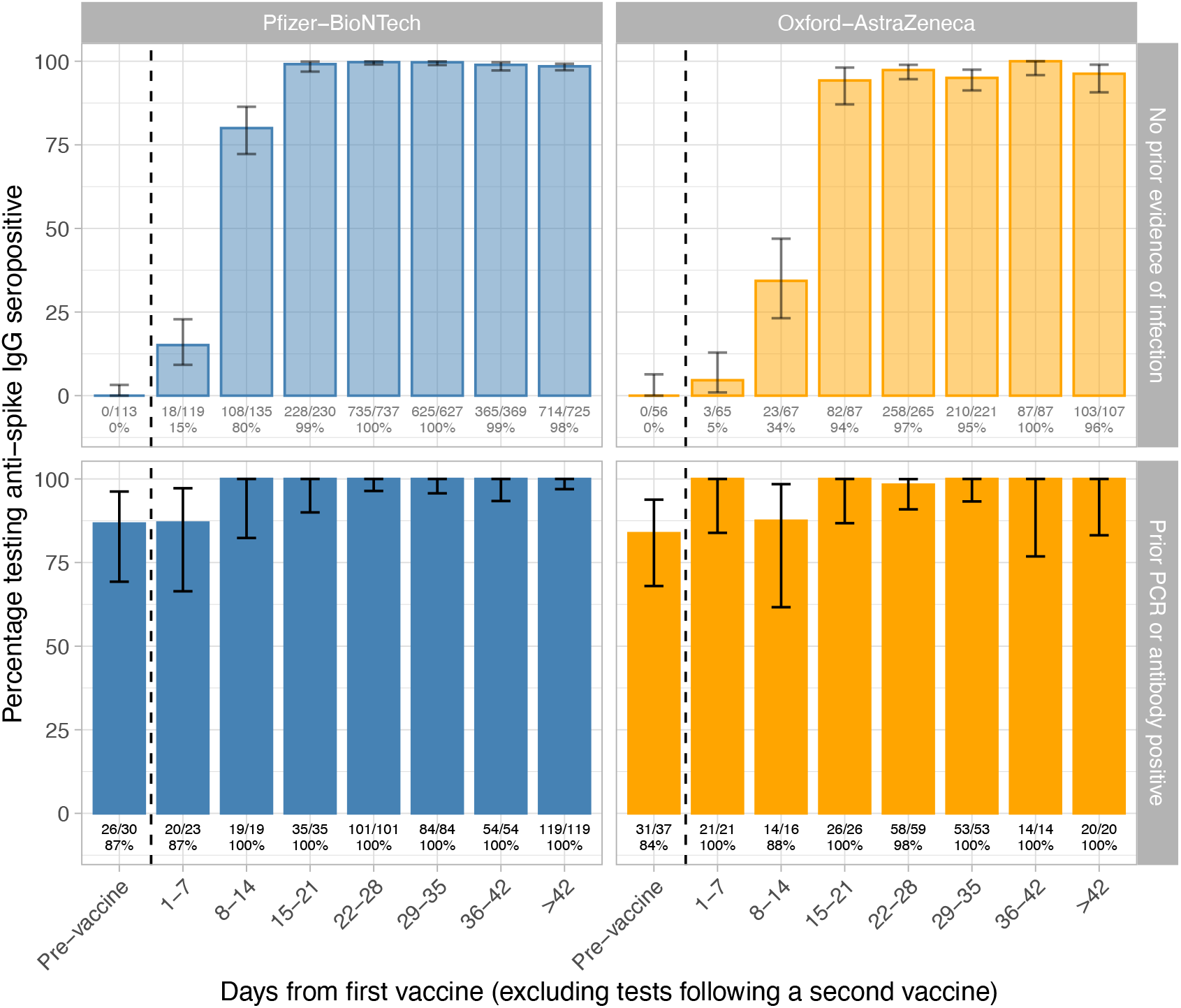
Anti-spike IgG positive results by days since first vaccination, by prior infection status and vaccine received. Tests performed after a second dose of vaccine are not included. The number of tests performed and positive and the resulting percentage is shown under each bar.

In a regression model of responses >14 days post-first vaccine and before second vaccine, previously infected HCWs were independently more likely to test seropositive (in part reflecting previously positive serology was one of the criteria for assessing previous infection; adjusted odds ratio, aOR 6.99, 95%CI 0.95-51.3, p=0.06) and older HCWs were less likely (aOR per 10 year older 0.66, 95%CI 0.51-0.86, p=0.002). There was no evidence of an effect of sex or ethnicity. Oxford-AstraZeneca vaccine recipients were less likely to seroconvert after their first vaccine dose versus Pfizer-BioNTech recipients (aOR 0.17, 95%CI 0.09-0.33, p<0.001). However, the absolute probability of seroconversion remained near 100% across most groups, except older HCWs receiving the Oxford-AstraZeneca vaccine, .e.g. a 60 year-old white female HCW had a 98.8% (95%CI 97.7-99.4%) chance of seroconversion with Pfizer-BioNTech and 93.2% (95%CI 89.0-95.9%) with Oxford-AstraZeneca post-first dose (Figure 2).

**Figure 2.**
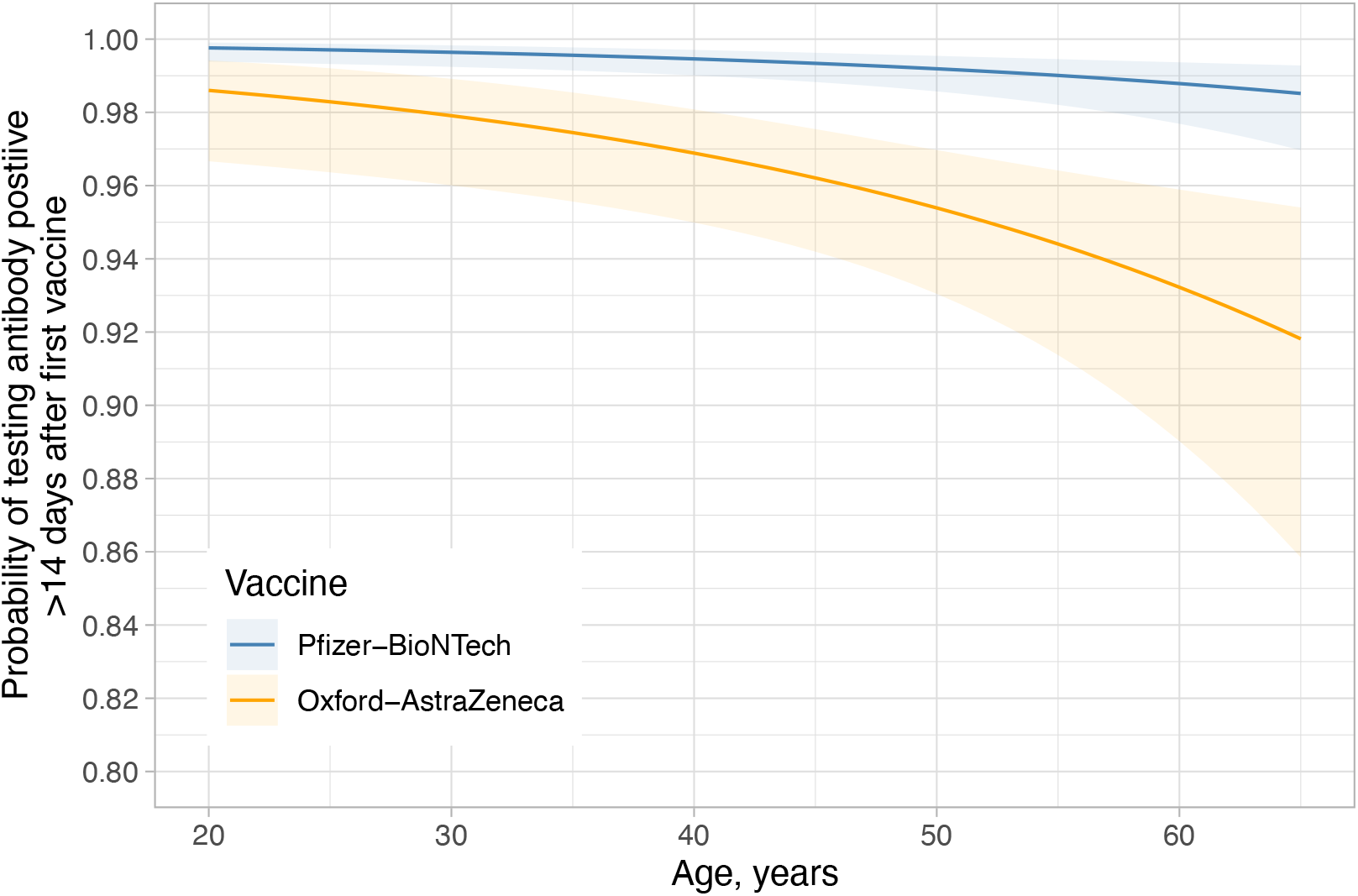
The relationship between vaccine, age and probability of testing anti-spike IgG seropositive >14 days post-first vaccination. Model predictions are shown using reference categories for sex and ethnicity (white, female, respectively) and in those without prior evidence of infection.

All 448 HCWs with an antibody test >14 days after their second Pfizer-BioNTech vaccine were seropositive. There were relatively few HCWs vaccinated twice with the Oxford-AstraZeneca vaccine, but all 22 assayed >14 days post-second dose were seropositive (Figure S2).

### Quantitative antibody readings before and after vaccination

Pre-vaccination quantitative antibody levels were available in 67 previously infected HCWs and 169 without evidence of prior infection; median (IQR) readings were 334 (103-1070) and 0.1 (0-1.4) AU/ml respectively. The median (IQR) time from first evidence of previous infection (first positive PCR or serological test) in those previously infected was 31 (0-246) days, with no evidence of association with antibody levels (Spearman’s rho=-0.09, p=0.45; Figure S3).

Quantitative vaccine readings rose during the 3 weeks post-first vaccination before plateauing (Figure 3). Those with previous infection developed substantially higher titres. In those receiving the Pfizer-BioNTech vaccine, the median (IQR) anti-spike IgG reading >21 days post-first vaccine dose was 1028 (564-1985) AU/ml without evidence of prior infection and 14,604 (7644-22,291) AU/ml with (Kruskal-Wallis p<0.001). Those receiving the AstraZeneca vaccine had lower titres compared to the Pfizer-BioNTech, without and with previous infection 435 (203-962) AU/ml and 10,095 (5354-17,096) AU/ml respectively (p<0.001 vs. Pfizer-BioNTech and within AstraZeneca). In previously uninfected HCWs, after Pfizer-BioNTech vaccination higher titres were seen in younger age groups (Figure 3C). Otherwise, there was no clear relationship between age and post-vaccine antibody readings.

**Figure 3.**
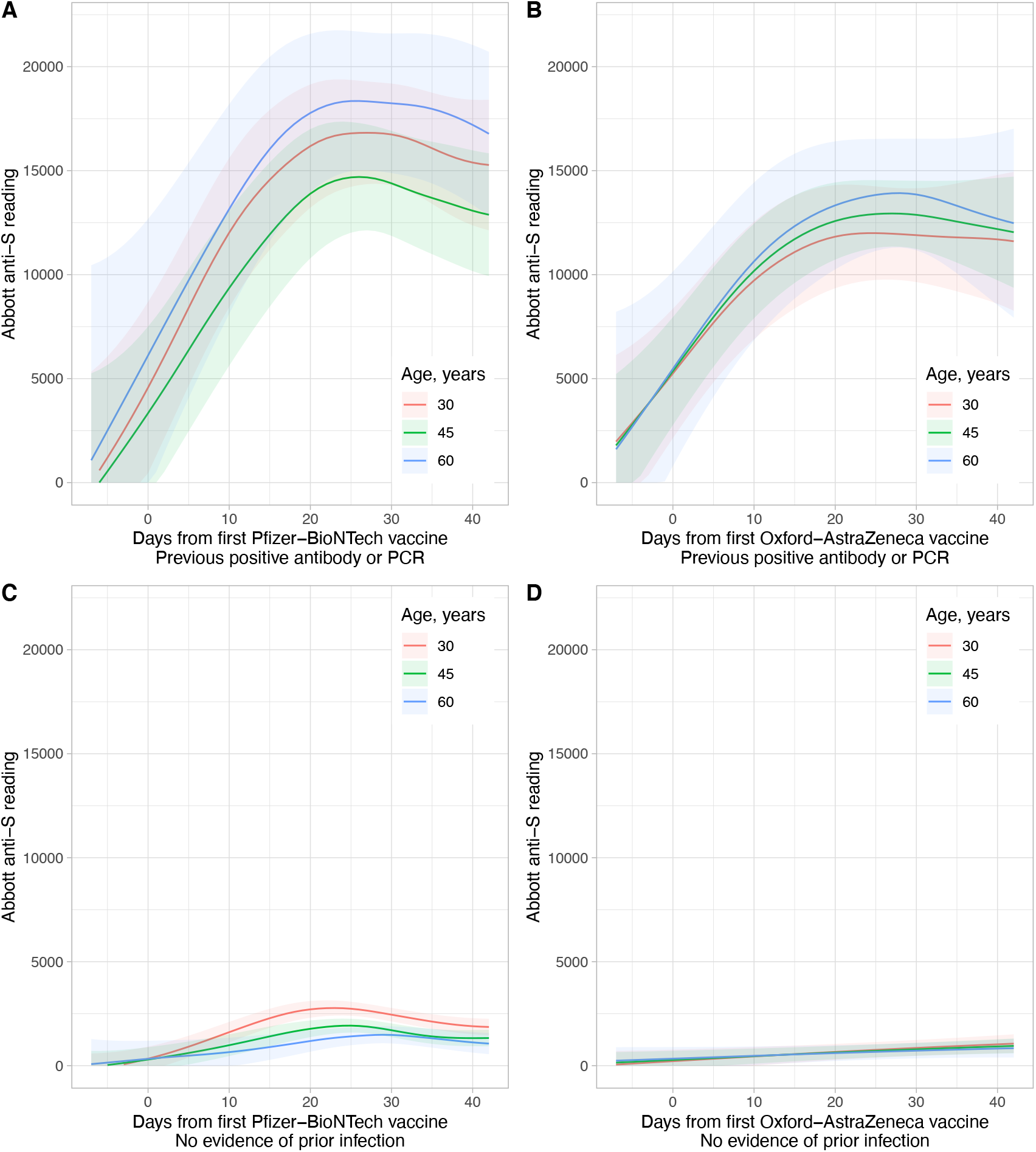
Modelled quantitative anti-spike IgG responses following first vaccination by vaccine and previous infection status. Panels A and B show responses in previously infected HCWs and panels C and D HCWs without evidence of previous infection. Panels A and C show data for those receiving Pfizer-BioNTech vaccine and panels B and D Oxford-AstraZeneca vaccine. Model predictions are shown at 3 example ages, 30, 45, and 60 years. The shaded ribbon shows the 95% confidence interval. Values are plotted from 7 days prior to vaccination to illustrate baseline values (models are fitted using data from 28 days prior to vaccination onwards).

In HCWs receiving a second Pfizer-BioNTech vaccine dose, antibodies were boosted in previously uninfected individuals with the highest levels in younger HCWs, but with some waning of responses from day 20-60 post-vaccination (Figure 4). Median (IQR) anti-spike IgG readings >21 days post-second vaccine dose were 10,058 (6408-15,582) AU/ml without evidence of previous infection and 18,047 (10,884-22,413) AU/ml with. Hence, anti-spike readings post-second vaccine in those without evidence of previous infection (Figure 4B) were similar to those seen after one vaccine in previously infected HCWs (Figure 3A/B).

**Figure 4.**
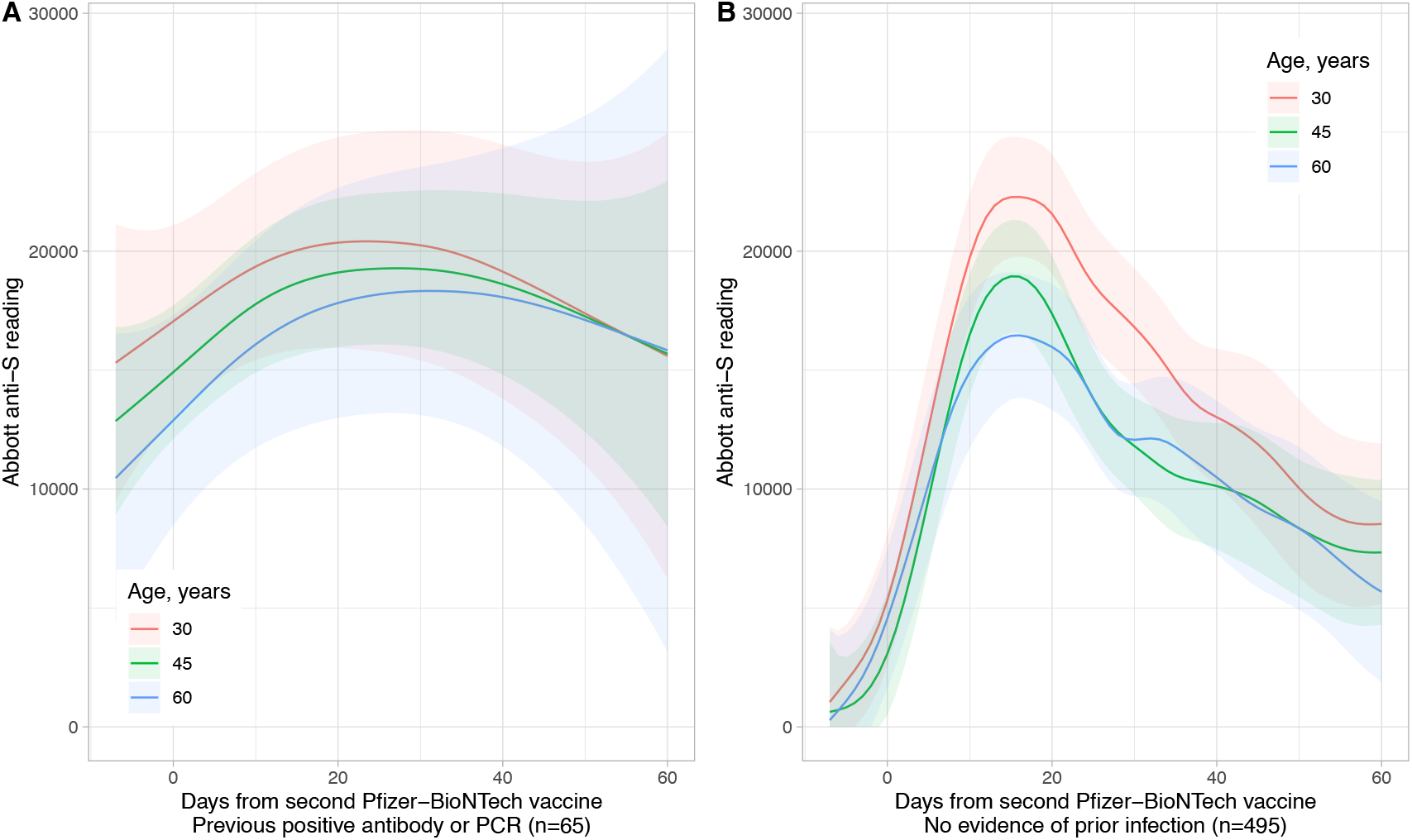
Modelled quantitative anti-spike IgG titres following second Pfizer-BioNTech vaccination by previous infection status. Panel A shows those who were previous infected (including those previously infected at baseline or testing PCR positive between vaccines) and panel B those who had no evidence of previous infection. Model predictions are shown at 3 example ages, 30, 45, and 60 years. The shaded ribbon shows the 95% confidence interval. Data were included in each model from 7 days before second vaccination to allow pre-vaccination levels to be fitted correctly.

Of 4069 HCWs who had an anti-spike IgG measurement >14 days after first vaccination, including those assessed after a second vaccination, only 9 HCWs had a subsequent positive PCR test after their antibody test (median [range] 48 [37-70] days post-first vaccine). Therefore, there was very limited power to detect any relationship between protection and quantitative antibody readings. The median (IQR) [range] maximum post-vaccine, pre-infection antibody measurement in those infected was 1062 (735-2497) [644-16,510] AU/ml and 1360 (580-4599) [0-40,000] AU/ml in those not (Kruskal-Wallis p=0.85).

## Discussion

In this cohort of >4000 HCWs, predominantly healthy adults of working-age, 98.9% developed a positive anti-spike IgG antibody test by >14 days post-first vaccine. HCWs in their 50s and 60s were less likely to seroconvert than younger HCWs, but absolute rates of seroconversion remained high in all groups. Fewer HCWs tested seropositive after a first dose of Oxford-AstraZeneca compared to Pfizer-BioNTech vaccine, with the difference more marked as age increased. Following a first dose, Pfizer-BioNTech vaccine resulted in higher antibody readings than Oxford-AstraZeneca vaccine in both those with and without previous infection, over 2-fold higher if not previously infected. All HCWs assessed >14 days post-second vaccine tested seropositive, although this included only 22 Oxford-AstraZeneca recipients.

The biggest determinant of the magnitude of quantitative antibody responses post-first vaccine dose was previous infection, with median readings >10-fold higher with previous infection for both vaccines compared to without previous infection. Antibody levels after prior infection and a single vaccine dose were similar to those achieved after two vaccine doses. Taken together with evidence that natural infection without vaccination offers similar protection from infection to two doses of vaccine,^17^ these data support prioritising uninfected individuals where vaccine sparing strategies are required. It may be possible to delay first vaccination in previously infected healthy individuals and one dose may be sufficient if there is robust serological evidence of previous infection.

We found lower rates of seroconversion in older HCWs (i.e. ∼60 years). The Pfizer-BioNTech trial reported neutralising titres and S1-binding antibody concentrations were higher in younger (18-55 years) vs. older (56-85 years) participants,^6^ however high seroconversion rates by S-binding antibodies were observed in Oxford-AstraZeneca trials involving older adults (≥65 years) after their first (97.3% [N=149, 95% CI: 93.3-99.3]) and second doses (100.0% [N=156, 95% CI: 97.7-100.0]).^8^

Although we observed differences in the proportion seroconverting and the magnitude of response following Pfizer-BioNTech and Oxford-AstraZeneca vaccines, this should not be taken alone as evidence that one vaccine is likely to be more efficacious than another. Anti-spike antibodies titres are associated with neutralising activity^1–3^, but the degree to which binary or qualitative anti-spike results are a surrogate from protection against infection, or other endpoints of interest such as hospitalisation, death or onward transmission, remains unclear. Our study was insufficiently powered to determine the relationship between antibody titres and protection, with only 9 HCWs infected after a post-vaccine antibody test. However, all 9 had positive antibody results with readings ranging from 644 to 16510 AU/ml, such that much of the range of positive anti-spike readings recorded in the study is not associated with total protection from infection. Although nearly all staff seroconverted post-first dose, we have previously shown single-dose vaccine effectiveness to be 67% in this period.^17^ This discrepancy between seroconversion and protection against reinfection, particularly post-first dose, is similar to that reported in Oxford-AstraZeneca trial participants; 98.5% seroconverted 28 days post-first dose, however pooled vaccine efficacy post-first dose was 76.0% (95%CI 59.3-86.9%).^18^ Seroconversion rates are not reported in the Pfizer-BioNTech trial. This discrepancy may in part be because the protective anti-spike threshold may differ from the assay positive/negative threshold, and also due to variation in protection mediated by other mechanisms. Further large-scale studies will be required to determine whether quantitative antibody levels can act as a surrogate marker for protection. Antibody testing post-vaccination may also play other roles. It may be useful to identify individuals in high risk groups who do not seroconvert^19^, who may remain at higher risk of infection, and may benefit from tailored advice around social contact, and, depending on underlying comorbidity, from further vaccination doses e.g. if immune reconstitution is expected.

Overall, we observed 80% vaccine uptake. Patterns broadly follow those seen more widely in the UK^20–23^ with lower uptake in HCWs from lower socioeconomic groups and of Black, south Asian and minority ethnic groups. This is particularly challenging as these groups are among those at highest risk of infection, independent of vaccination status.^17^ OUH is actively targeting under-vaccinated staff groups with specific advertising, outreach and mobile vaccination facilities.

Study limitations include the use of a single assay to quantify post-vaccine anti-spike antibody levels, however as it is commercially-available and well calibrated (Figure S1), results should be generalisable. Our focus on a defined group, i.e. HCWs, is both a strength and a weakness, and we are not able to assess variations in vaccine response in children or those >65 years. Additional data on post-vaccine antibody responses in older individuals, at highest risk of adverse outcomes from SARS-CoV-2 infection, is particularly important. Our cohort was also 76% female and predominantly of white ethnicity. We did not assess neutralising antibodies or T cell responses, both of which reflect vaccine response and may vary by vaccine. Further work will be required to determine the duration of antibody responses.

In summary, vaccination leads to detectable anti-spike antibodies in nearly all healthy adult HCWs. Markedly higher responses to vaccine are seen after previous infection; single dose or delayed vaccination could be considered where vaccine sparing is needed in healthy individuals with robust evidence of previous infection. Some caution is required with antibody result interpretation and any subsequent behaviour change, as despite good protection from vaccination, seroconversion with high antibody levels still does not afford absolute protection from infection. Large-scale studies will be required to assess how protection from infection varies by antibody titre.

## Data Availability

The datasets analysed during the current study are not publicly available as they contain personal data but are available from the Infections in Oxfordshire Research Database (https://oxfordbrc.nihr.ac.uk/research-themes-overview/antimicrobial-resistance-and-modernising-microbiology/infections-in-oxfordshire-research-database-iord/), subject to an application and research proposal meeting the ethical and governance requirements of the Database.

## Acknowledgements

We would like to thank all OUH staff who participated in the staff testing program, and the staff and medical students who ran the program. This work uses data provided by healthcare workers and collected by the UK’s National Health Service as part of their care and support. We thank all the people of Oxfordshire who contribute to the Infections in Oxfordshire Research Database. Research Database Team: L Butcher, H Boseley, C Crichton, DW Crook, DW Eyre, O Freeman, J Gearing (community), R Harrington, K Jeffery, M Landray, A Pal, TEA Peto, TP Quan, J Robinson (community), J Sellors, B Shine, AS Walker, D Waller. Patient and Public Panel: G Blower, C Mancey, P McLoughlin, B Nichols.

## Author contributions

Conceptualisation: DWE, SFL, DWC, TEAP, TMW, NEW, PCM, KBP, ASW, KJ

Methodology: DWE, JW, KBP, ASW

Formal analysis: DWE

Investigation: SC, TJ, AJ, GJ, DOD, AH, SBH, BDM, DE

Writing – Original Draft: DWE, SFL

Writing – Review & Editing: All authors

Visualisation: DWE

Supervision: DWE, EYJ, DIS, DE, SH, DWC, TEAP, TMW, ASW,KJ

Project administration: SH

