## Supplement for "Quantitative SARS-CoV-2 anti-spike responses to Pfizer-BioNTech and Oxford-AstraZeneca vaccines by previous infection status"

### Supplementary methods

#### PCR assays

RT-PCR was performed using the Public Health England SARS-CoV-2 assay (targeting the RdRp gene), one of five commercial assays: Abbott RealTime (targeting RdRp and N genes; Abbott, Maidenhead, UK), Altona RealStar (targeting E and S genes; Altona Diagnostics, Liverpool, UK), Cepheid Xpert® Xpress SARS-CoV-2 (targeting N2 and E; Cepheid, California, USA), BioFire® Respiratory 2.1 (RP2.1) panel with SARS-CoV-2 (targeting ORF1ab and ORF8; Biofire diagnostics, Utah, USA), Thermo Fisher TaqPath assay (targeting S and N genes, and ORF1ab; Thermo Fisher, Abingdon, UK) or using the ABI 7500 platform (Thermo Fisher, Abingdon, UK) with the US Centers for Disease Control and Prevention Diagnostic Panel of two probes targeting the N gene.

PCR-positive results from community-based symptomatic testing of Oxford University Hospitals (OUH) healthcare workers (HCWs) forwarded by public health agencies were also included (Thermo Fisher TaqPath assay).

#### Serological assays

The manufacturer's reported sensitivity of the Abbott SARS-CoV-2 IgG II Quant antibody test is 157/158 (99.4%, 95%CI 96.5-99.9%) ≥15 days after symptom onset in individuals with PCR-confirmed SARS-CoV-2 infection. Reported specificity is 1999/2008 (99.6%, 95%CI 99.2-99.8%) (assay instructions for use).

#### Vaccinations

Staff vaccinations provided by the hospital were recorded on an electronic database linked to testing data; staff were also able to supply details of vaccinations from other providers when requesting a symptomatic or asymptomatic PCR test.

#### Statistical models

Analyses were undertaken using R (v.4.04) and the mgcv (v.1.8-34) and splines (v3.6.2) libraries. We used multivariable logistic regression to identify predictors of a positive anti-spike antibody result at any time ≥15 days post-first vaccination (but before a second vaccination), considering the vaccine given, previous infection status, age, sex, and ethnicity. We allowed for non-linear effects of age using natural cubic splines with up to 5 default spaced knots, choosing the best fitting model using Akaike information criterion

We modelled quantitative antibody titres by day since post-first vaccination using generalised additive models, adjusting for age. Separate models were fitted by previous infection status and for each of the two vaccines. To allow for estimation of baseline pre-vaccine antibody levels, antibody readings from up to 28 days prior to first vaccination were included. We allowed for non-linear main effects and interactions for time since vaccination and age using tensor product smoothers. We allowed for repeat measurements in the same individual by including a random effect smoother. Example code for running the analysis is provided below, where the basis dimensions k1 and k2 are chosen based on optimising model fit:

```
m = bam(abbott_spike_reading ~ te(days_from_vaccine1, age, bs="tp",
k=c(k1,k2))+ s(user_id, bs="re"), method = "REML", data=df)
```

Similar models were fitted following a second vaccine dose for those receiving the Pfizer-BioNTech vaccine. Insufficient data were available after a second Oxford-AstraZeneca vaccine to fit models. Results from up to 7 days before second vaccine were included to allow for estimation of levels at the time of second vaccination.

### Supplementary figures

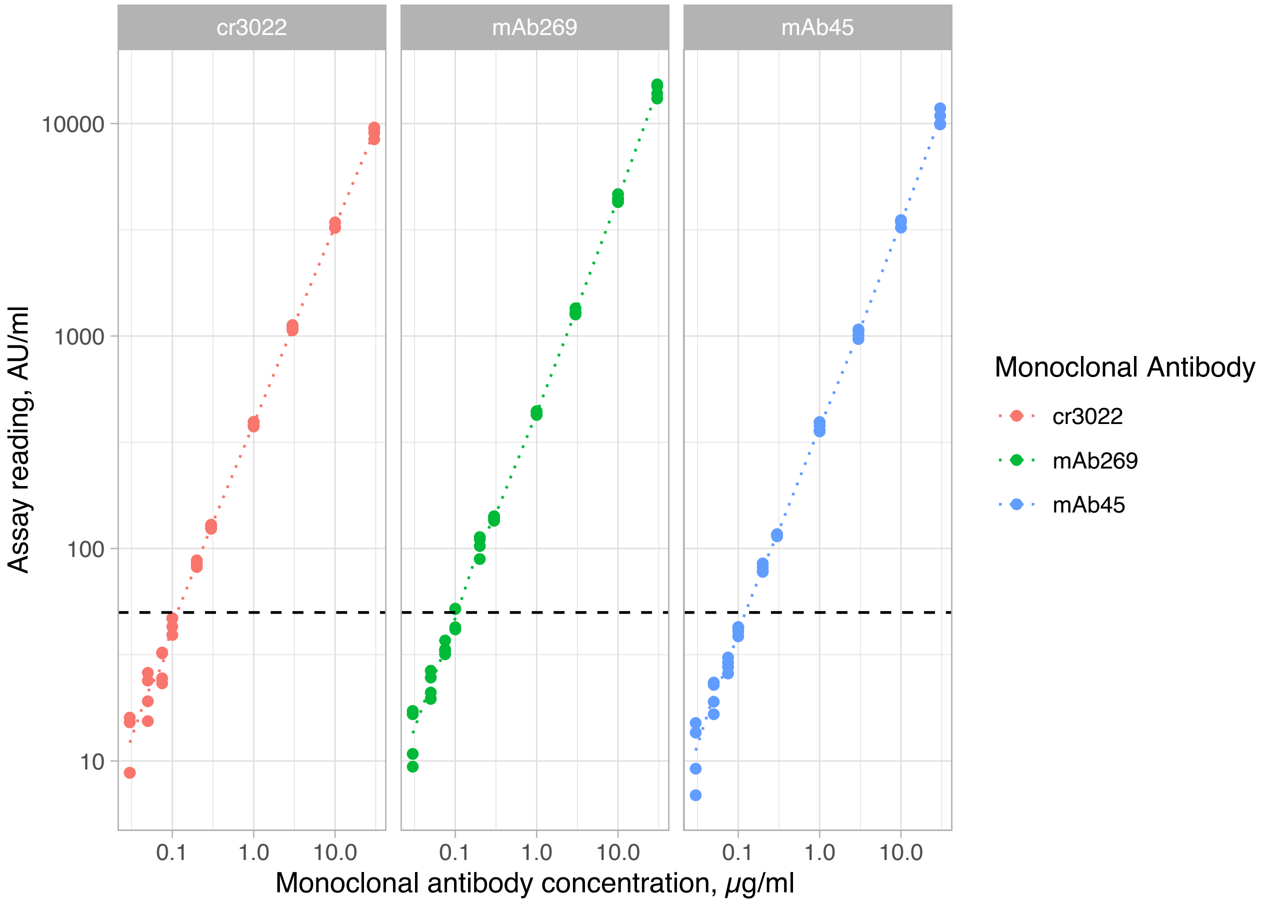


**Figure S1. Abbott SARS-CoV-2 IgG II Quant antibody test calibration**. A linear response is seen between the concentration of 3 SARS-CoV-2 anti-spike monoclonal antibodies (cr3022, mAb269, mAb45) and the assay reading. Both axes are shown on a log10 scale. The horizontal dashed line indicates the assay cut-off of ≥50 AU/ml. The dotted line for each assay joins the mean values at each dilution.

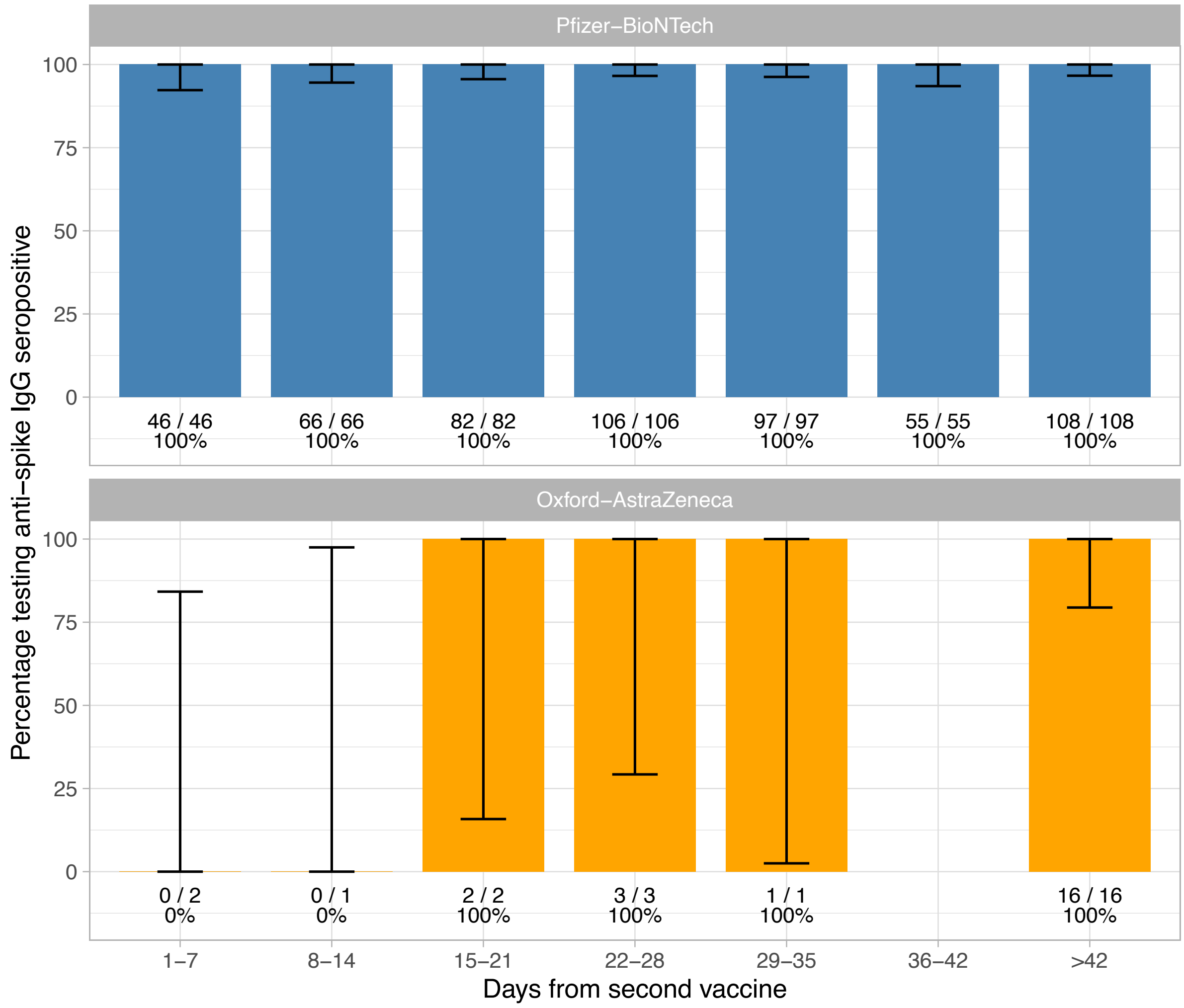


**Figure S2. Anti-spike IgG antibody status by vaccine and days since second vaccination**.

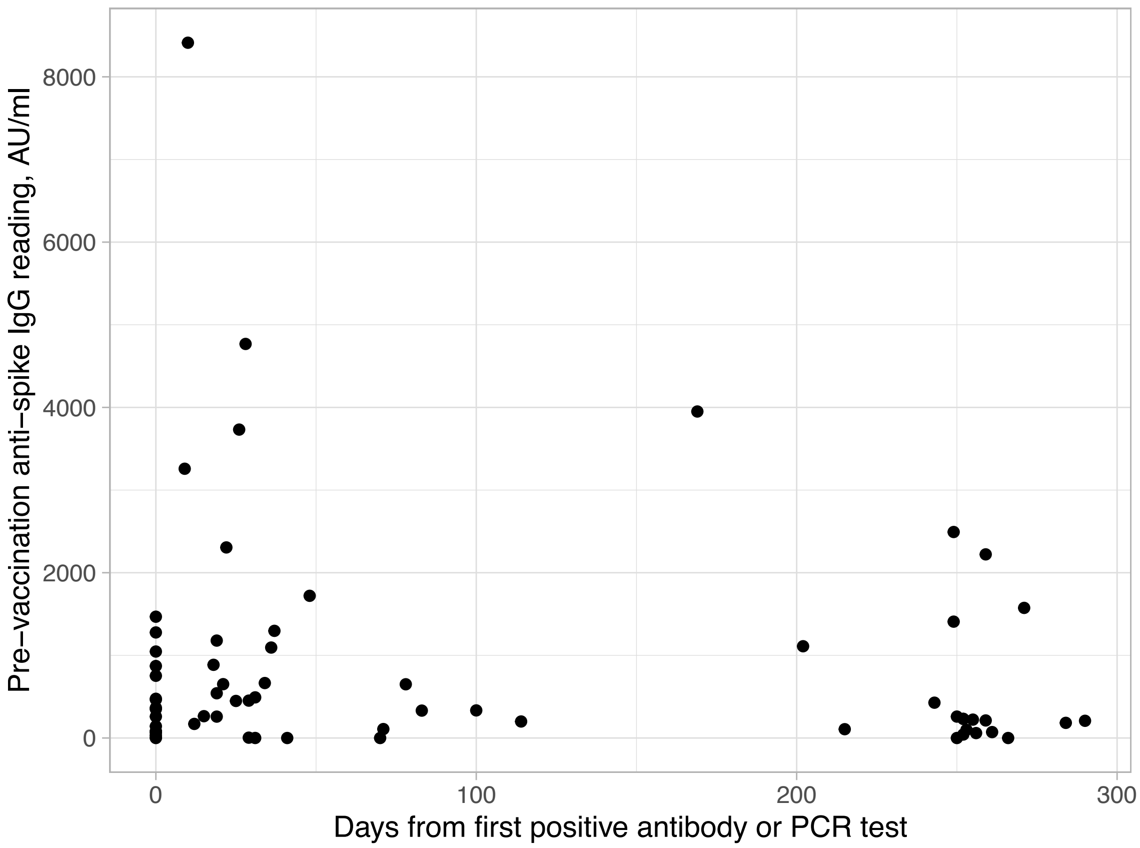


**Figure S3. Pre-vaccination anti-spike antibody levels in 67 previously infected HCWs**.
